# Prediction of Coronavirus Disease (covid-19) Evolution in USA with the Model Based on the Eyring’s Rate Process Theory and Free Volume Concept

**DOI:** 10.1101/2020.04.16.20068692

**Authors:** Tian Hao

## Abstract

A modification arguing that the human movement energy may change with time is made on our previous infectious disease model, in which infectious disease transmission is considered as a sequential chemical reaction and reaction rate constants obey the Eyring’s rate process theory and free volume concept. The modified model is employed to fit current covid-19 outbreak data in USA and to make predictions on the numbers of the infected, the removed and the death in the foreseeable future. Excellent fitting curves and regression quality are obtained, indicating that the model is working and the predictions may be close to reality. Our work could provide some ideas on what we may expect in the future and how we can prepare accordingly for this difficult period.

## I. INTRODUCTION

During this global pandemic outbreak of coronavirus 2019 (*covid* – 19), USA becomes the number one in term of how many people are infected. What is going to happen next and how many people may be infected and may die become an emergent question for policy makers to make proper mitigation plans. Mathematical modeling and analysis of infectious disease transmissions^1–8^ have been utilized to make predictions. Precise prediction remains challenging due to randomness of human interactions and unpredictability of virus growth patterns. Human mobility and virus transmissions, however, should follow basic physical and chemical laws. Two very powerful theories in physics and chemistry fields are the Eyring’s rate process theory and the free volume concept. The Eyring’s rate process theory^9^ argues that every physical or chemical phenomenon is a rate controlled process, while the free volume concept^10–14^ argues that the transmission speed is also dependent on the available free volume. Many seemingly unrelated systems or phenomena can be successfully described with these two theories, such as glass liquids^13^, colloids and polymers^15,16^, granules^17–19^, electrical and proton conductivity^20,21^, superconductivity^22^, and Hall Effect^23^, etc. The infectious disease transmission phenomenon, a very complicated macroscopic process, could be properly analyzed with these two theories, too. Attempts were made to integrate these two theories together for modeling infectious disease transmissions under an assumption that an infectious disease transmission is a sequential chemical reaction^1^. Focus was placed on analyzing covid-19 outbreak in China for validating the newly formulated model and making predictions on peak time and peak infected.

In this article, an infectious disease is still considered as a sequential chemical reaction by following the popular SIR (*susceptible, infectious, and removed*) and SEIR (*susceptible, exposed, infectious, and removed*) compartment categorization methods proposed in the literature^2–8^. For better fitting data, modification is made on our previous model^1^ by introducing an idea that the energy for human individuals to transmit diseases is time dependent, which is in line with other systems like granular powder under tapping process where the energy of particles is time dependent, too^18^. The modified model is used to analyze covid-19 transmission in USA and make predictions on potential infections and death toll.

## II. THEORY

According to the model proposed previously^1^, the whole infection disease transmission process can be expressed as below:

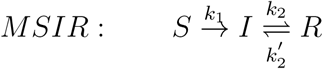

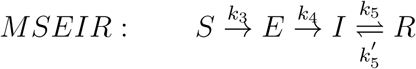

where *S, E, I*, and *R* represents the fractions or concentrations of the susceptible, the exposed, the infected, and the removed in the sequential chemical reaction. The difference between MSIR (modified Susceptible, Infectious, and Removed) and MSEIR (modified Susceptible, Exposed, Infectious, and Removed) models is that MSIR model assumes that the susceptible will directly transform into the infected, while MSEIR model assumes that there is an intermediate state “exposed”. *k*_1_, *k*_2_, 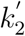, *k*_3_, *k*_4_, *k*_5_, 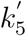 are chemical reaction rate expressed in two chemical reactions shown above. Once these parameters are known, the fractions of *S, E, I, R* can be predicted. For the first step reaction in MSIR model, I will follow th same approach used previously, i.e. two steps are involved during the process from ”*S*” to ”*I*”: human individual movement and virus particle movement. Human movement is an athermal stochastic random process. For athermal granular powder under a tapping process, we have demonstrated that they behave like thermal systems and follow the stretched exponential pattern in term of tap density changing with the number of taps^17^. Our approach is mainly based on Theodor F*ö*rster’s theory^24–27^ that deals with the energy transfer from donors to random distributed acceptors. Other people’s research work has shown that human collective motion and individual walking patterns of animals behave like thermal systems and follow Boltzmann distribution, though the term of temperature needs to be defined differently in these athermal systems^28,29^. For the process transformed from the susceptible to the exposed, human movements may play a major role and the reaction rate of this process, *k*_3_ in MSEIR model, may be expressed as^17^:

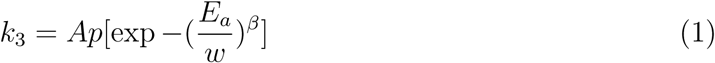

where *A* and *p* are constant, *β* is the stretched exponential parameter of a value between 0 and 1, *E_a_* is the energy for a human to attain for moving around during transmission period, and *w* is the basic/unit energy that a person may need during a normal circumstance, which is identical to the product of the Boltzmann constant and the temperature. Following the similar treatment method on powder particles^17^, we may assume that *E_a_* should be proportion to time and thus write:

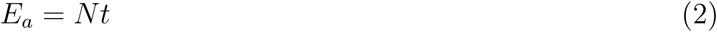

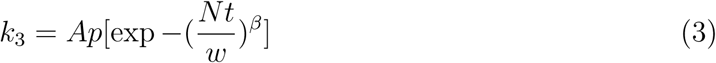

where *N* is a constant and *t* is the time. In previous article, *E_a_* was considered as a constant, independent of time. After an individual is exposed, the transmission of virus particles from one person to another will make an “exposed” person become “infected”. The transmission rate will be dependent on how fast these virus particles will travel and how large the free volume is available for virus particles to travel. It can be analogous to the viscosity or conductivity of an entity that has been addressed in many systems in my previous articles^13,15,20,21,30^ with free volume estimated using inter-particle spacing concept^15,31^. The chemical reaction rate should be proportional to the “viscosity” of this entity, then *k*_4_ can be written as^30^:

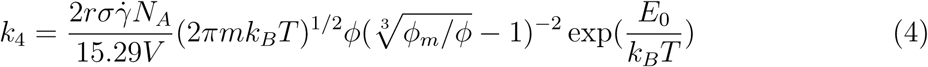

where *V* is the volume in consideration, *r* is the radius of a virus particle, *σ* is the shear stress applied when virus particles transmit from one place to another, 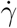 is the shear rate, *N_A_* is Avogadro number, *m* is the mass of a virus particle, *k_B_* is Boltzmann constant, *T* is the temperature, *ϕ* is the volume fraction of virus particles in the volume *V*, *ϕ_m_* is the maximum packing fraction of virus particles, and *E*_0_ is the energy barrier for virus particles.

In MSIR model, we assume that during the transmission process from the susceptible to the infected, both human movement and virus particle transmission are involved and the “exposed” is only an transient state. According to the transient state theory of chemical reaction^9^, we may easily obtain:

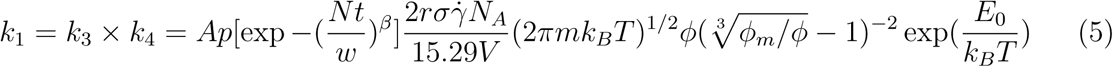

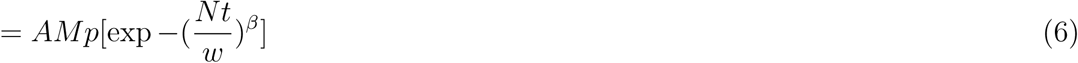

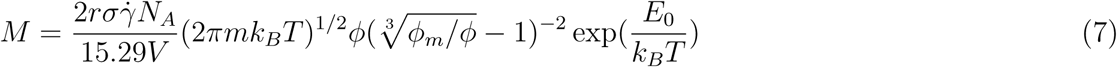

Eq. (1–7) indicates that infectious disease transmission is a complicated process, and is dependent on many factors like human movement energy barrier, the particle size and volume fraction of virus particles, the mass of a virus particle, temperature, and volume in consideration. Smaller volume leads to lower transmission rate, and isolation definitely is a good method to preventing virus from spreading.

For a sequential chemical reaction, the fraction of each reactant can be expressed with a series of differential equations^32^. For MSIR model, we may write:

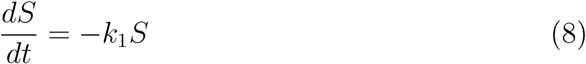

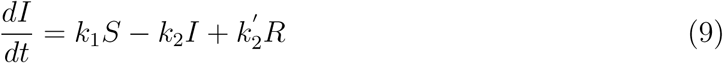

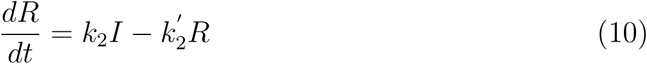

For MSEIR model, we may write:

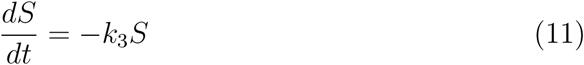

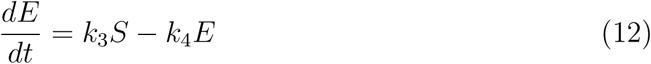

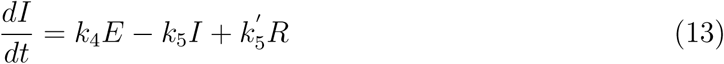

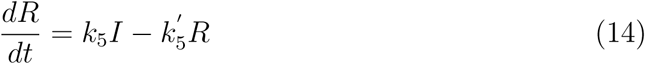

Assume that the initial fraction of the susceptible is *S*_0_ and *Nt* is always smaller than *w*, so *exp*(*−Nt/w*) ≅ 1 *−* (*Nt/w*)*^β^*. We may easily obtain:

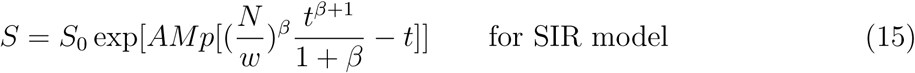

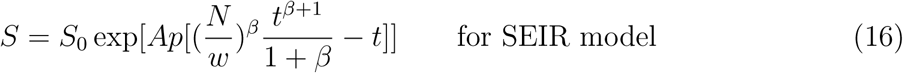

Since the contribution from 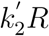 to the infected is negligible based on the fact that the recovered may gain immunity from the disease and the fraction of the recovered is relatively small at early stages, the first step reaction product, *E* and *I* may be written:

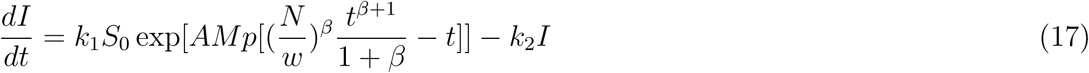

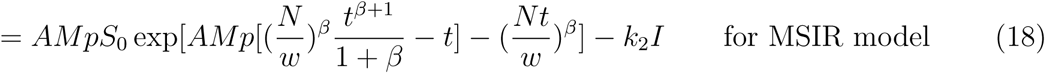

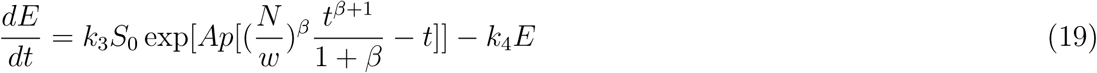

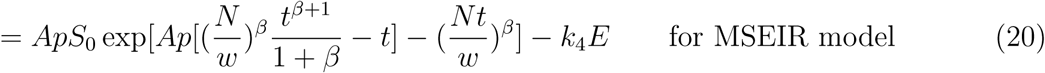

Both equations above are first-order differential equations of standard form:

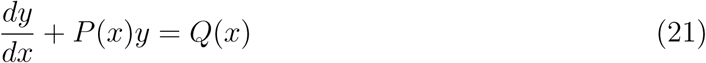

which has a standard solution as:

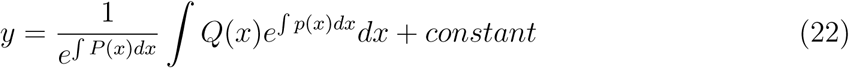

Using exp *x* ≅ 1 + *x* when *x* <1, we therefore obtain:

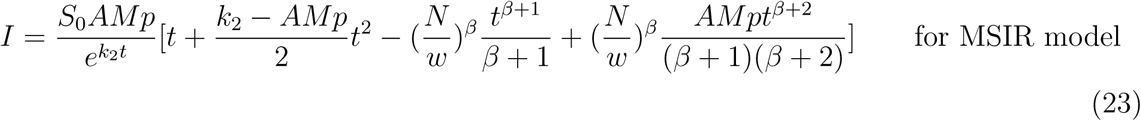

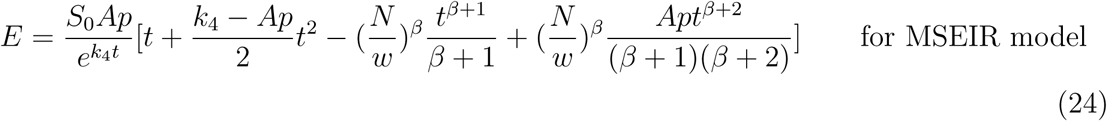

Similarly, ignoring the contribution from 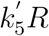, we may obtain *I* in MSEIR as shown below:

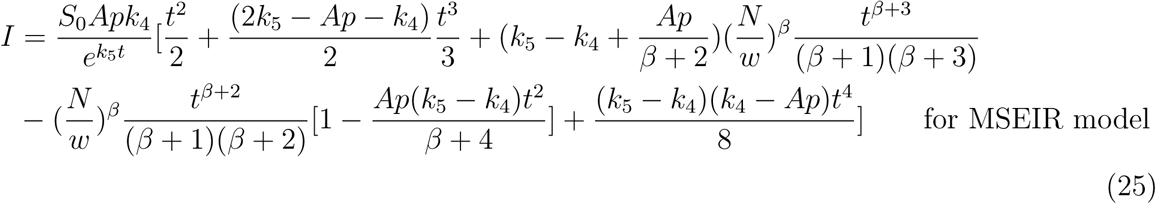

Assuming that *S* + *I* + *R* = *S*_0_ for MSIR model and *S* + *E* + *I* + *R* = *S*_0_ for MSEIR model, we can obtain:

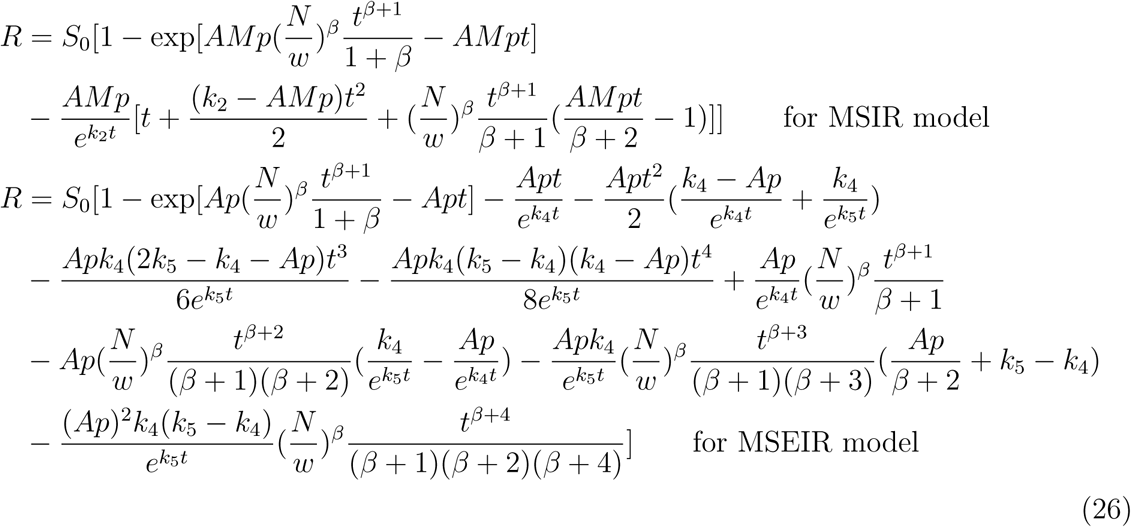

Both *I* and *E* should have a peak value that can be simply determined by differentiating Eq. 24 and Eq. 25 against time. If there is no exact analytical solution, we may determine approximate peak values after equations are plotted out.

## III. RESULTS

The fractions of the susceptible, exposed, infected, and recovered are functions of time, virus particle volume fraction, and environment temperature. The trends between these parameters had been graphed previously^1^ and shouldn’t be impacted by the modification of human movement energy term. Both the exposed and infected would peak at certain time, dramatically increase with virus particle volume fraction, and decrease with temperature increase. Please refer to my previous article for further information. Focus in this section will be placed on how the infected changes with other critical parameters like *β* and if these equations can be used to fit current data and make predictions.

The infected against both time and the stretched exponential parameter *β* are plotted in Figure 1. The infected peaks with time and increases with *β*. The parameter *β* basically enlarges peak heights, implying that when *β* is large, more peoples are infected. The physical meaning of *β*, according to Phillips^33,34^, is as below: *β* = 3/5 for intrinsic molecular level short range interactions, *β* = 3/7 for intrinsic long range coulomb interactions, and *β* = 2/3 for extrinsic interactions. With the increase of *β*, more interaction between the entity is expected, i.e. more infections, which seems to be logical in term of how infection transmission evolves.

**Figure 1:**
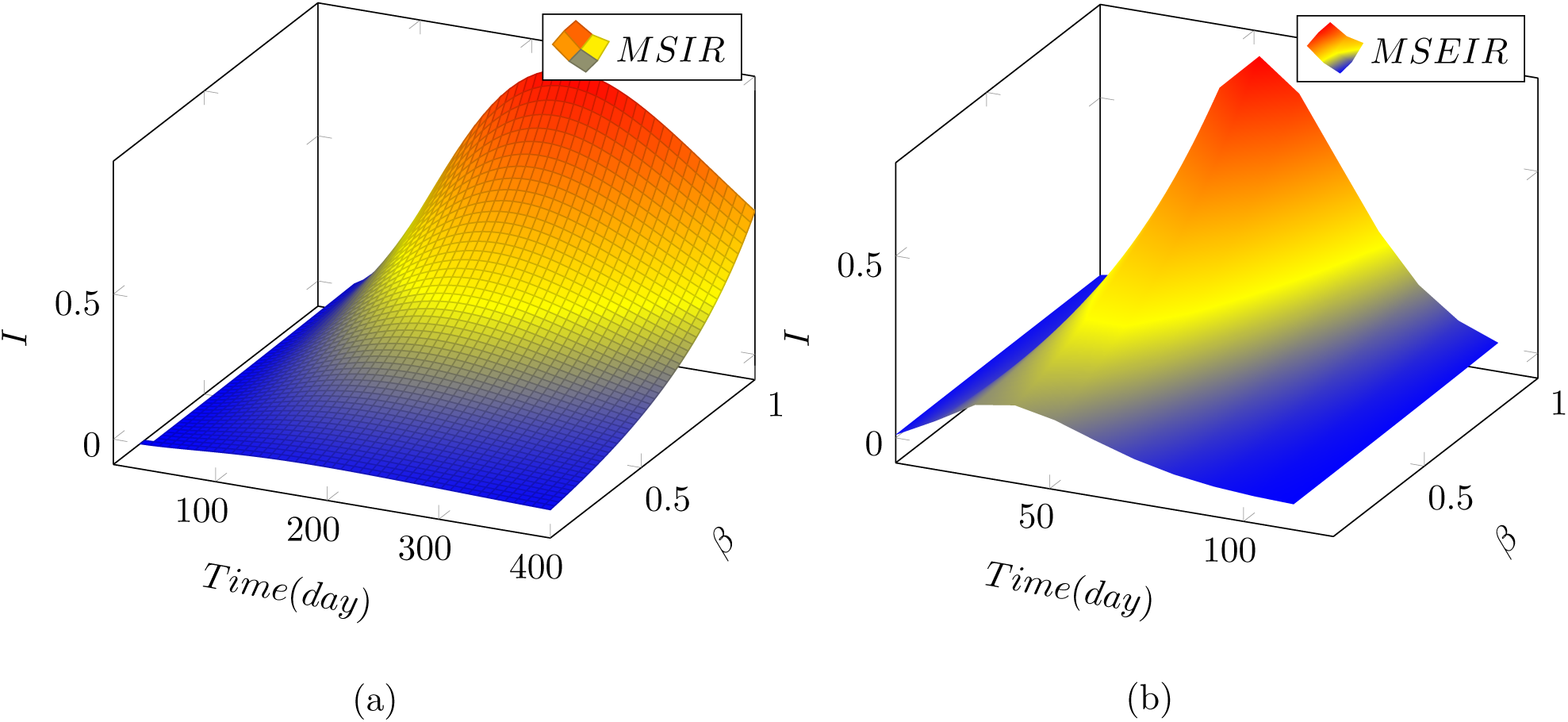
The fraction of the infected are plotted against both time and *β*. (*a*) predicted with MSIR model, and (*b*) predicted with MSEIR model

The rate constant could play a critical role in infection transmissions. The parameters *k*_1_ and *k*_3_ are replaced with other terms in equations and the remaining parameters like *k*_2_, *k*_4_ and *k*_5_ will be focused. Figure 2 shows the infected against both time and the parameter *k*_2_ for MSIR and the parameter *k*_4_ for MSEIR. The infection peak against time was not showing up in MSIR graph, probably due to unreasonable variations of *k*_2_. Peaks are shown up in Figure 1 when a fixed value of *k*_2_ is assigned. An infection peak against *k*_2_ is observed, indicating that the recovery rate is important in disease transmission, which may flatten the infection at very early stages. For MSEIR model shown in Figure 2 (b), high infection rate indicated by *k*_4_ means that the infection peaks at very early time and disappears quickly afterwards, possibly due to the fact that a large number of people are infected and “herd immunity” may be generated to stop further spreading.

**Figure 2:**
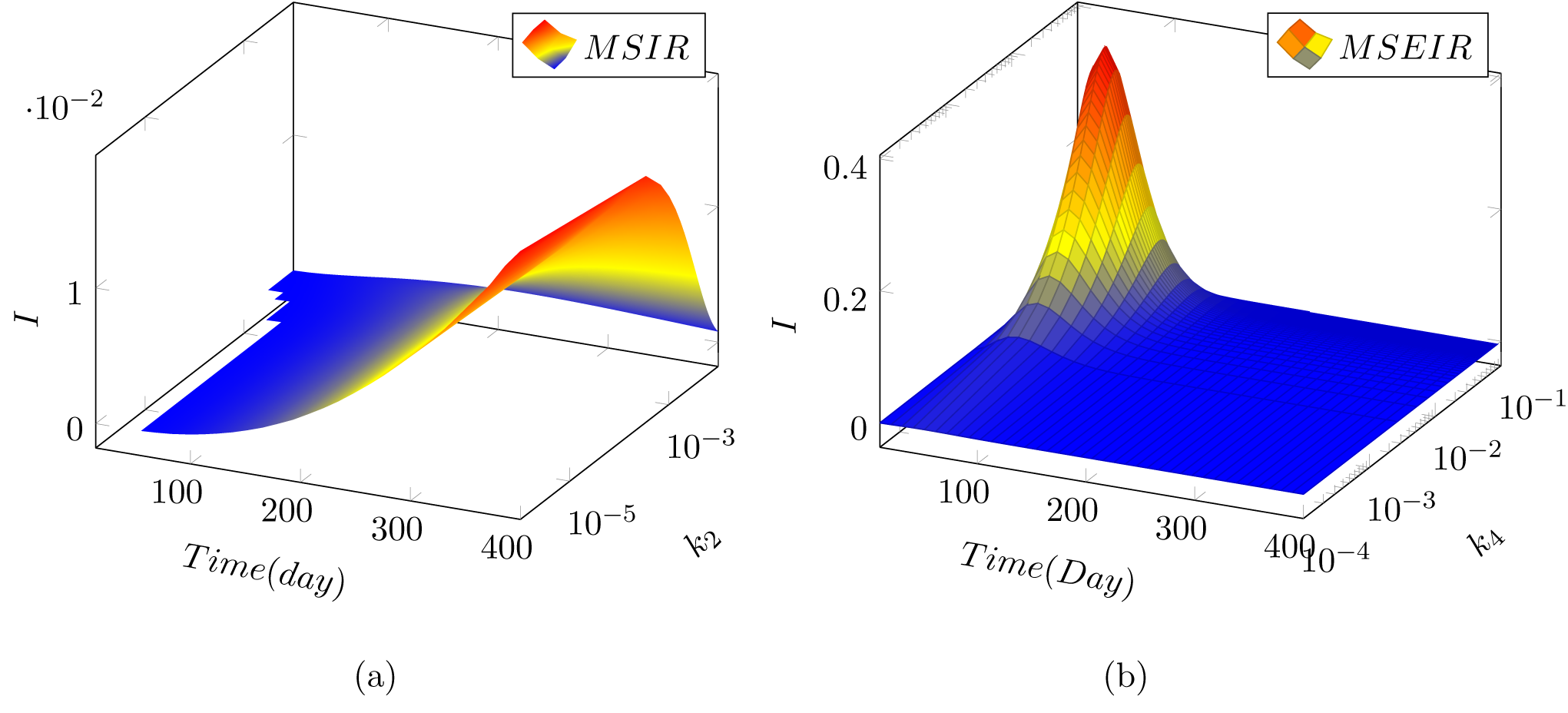
The fraction of the infected, (*a*) predicted with MSIR model for both time and *k*_2_, and (*b*) predicted with MSEIR model for both time and *k*_4_

Another two parameters in MSEIR model are AMp and *k*_5_, the impact of these two on the infected is shown in Figure 3. Similar to *β*, AMp enlarges or amplifies infection peak heights. Increase of AMp may have more people infected. The impact of *k*_5_ on the infected is similar to that of *k*_4_ shown in Figure 2. The infected peaks at a early time when more people can be removed from the system, including the recovered and death.

**Figure 3:**
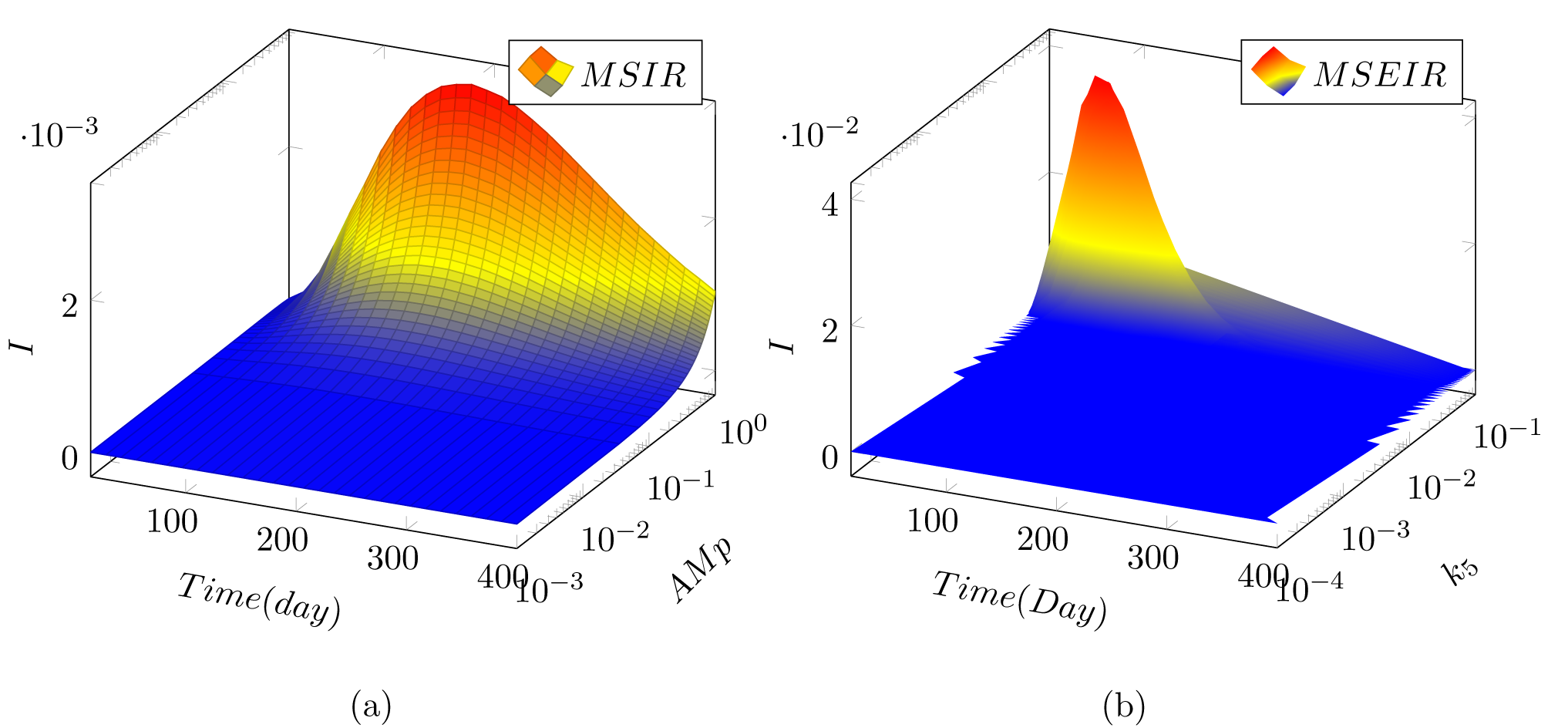
The fraction of the infected are plotted against both time and AMp (a) and *k*_5_ (b) for MSEIR model

If the equations created are correct, they should be able to fit data and make predictions on what is going to happen next. Figure 4 shows the fraction of the infected in USA from March 12 to April 28, 2020 and fitted with both MSIR and MSEIR models. The *R*^2^ of both fittings are larger than 0.99 as demonstrated in Figure 4(a). Same equations with same fitting parameters are plotted again in a larger scale and shown in Figure 4 (b). The first numbers showing on peak points are the peaking days from March 12, 2020, and the second numbers are the peak fractions. MSIR model predicts that the infected in USA may peak 115 days from March 12, about on July 7, 2020, while MSEIR predicts that the infected in USA may peak 64 days from March 12, about on May 16, 2020. The peak infected is also quite different: 1.98 million predicted with MSIR and 1.08 million predicted with MSEIR. *β*=0.2 is used for both fitting process. Such a small *β* value indicates that severe interpersonal transmission is not happening yet.

**Figure 4:**
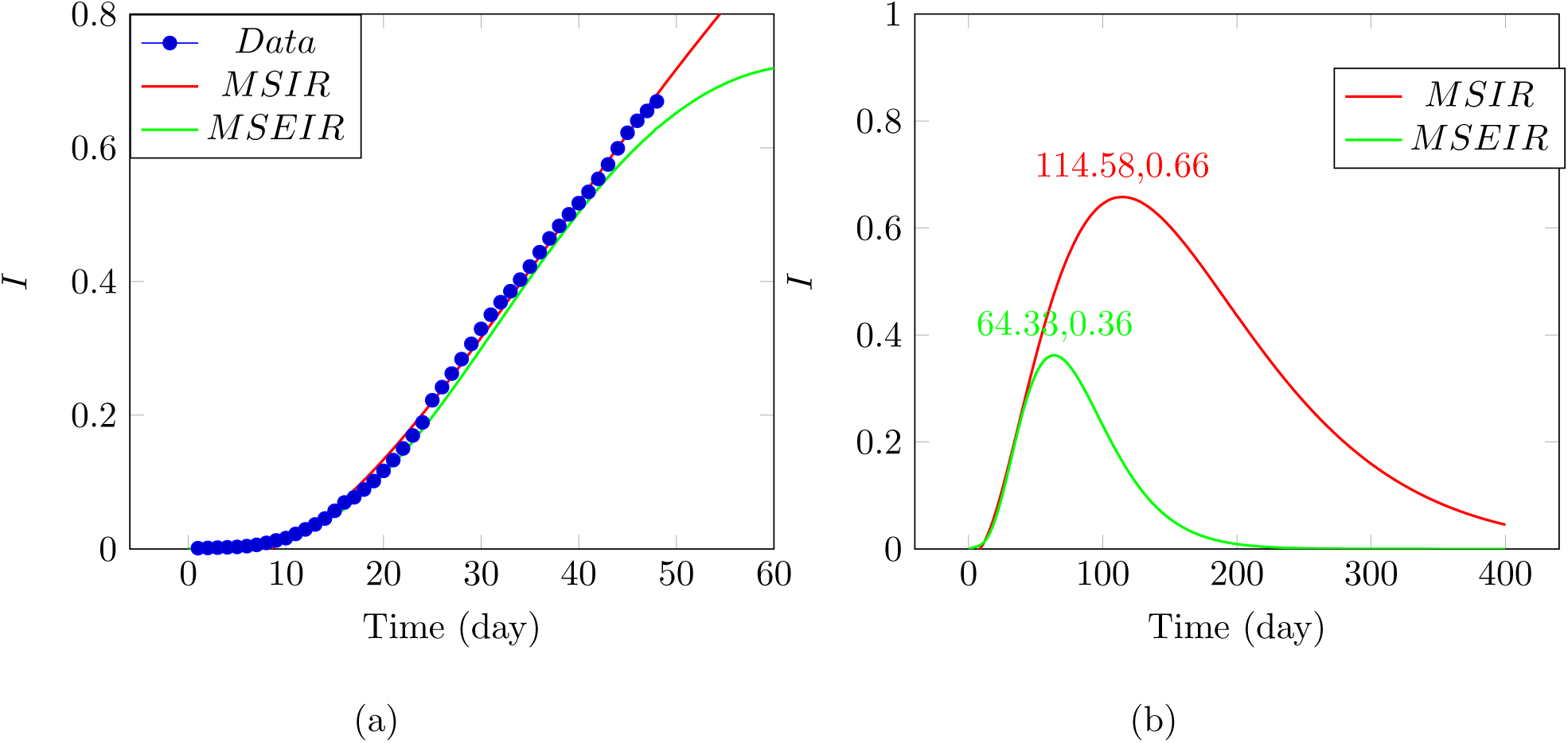
The fraction of the infected calculated from coronavirus disease (Covid-19) data https://en.wikipedia.org/wiki/2020_coronavirus_pandemic_in_the_United_States. These data are collected from the official reports from state health officials between March 12 to April 28, 2020, and fitted with MSIR and MSEIR models. The number of the infected individuals is divided by a population size 1.5 million to obtain the fraction. *R*^2^ for both regressions are larger than 0.99.

Another important information is the removed, including both the recovered and the death. Figure 5 shows two sets of data, one is the recovered alone and another includes both the recovered and the death. Both sets of data are fitted with MSIR and MSEIR models. Again, the fitting is very good with *R*^2^ larger than 0.99, though different fitting parameters are used for the recovered and the recovered plus the death. In both fitting processes *β*=0.01 is used, implying a very weak interpersonal interaction is found and there is no “herd immunity” happening. The large scale graphs calculated with the same fitting parameters are shown in Figure 6 for both the recovered alone (a) and the recovered plus the death (b). A huge number of people is projected to recover in the future. However, we’d better separate the recovered and the death from the removed. Eq. 26 is thus used to fit the death only data and shown in Figure 7 (a) for a small scale and (b) for a large scale. According to MSIR model, the death toll could reach 180,000 for 1.98 million infected at the peak time, about 9.06 % death rate; according to MSEIR model, the death toll could reach 80,000 for 1.08 million infected at the peak time, about 7.37 % death rate. The death rates predicted with MSIR and MSEIR models are higher than current death rate in USA, about 5.3 %, which actually increases with time.

**Figure 5:**
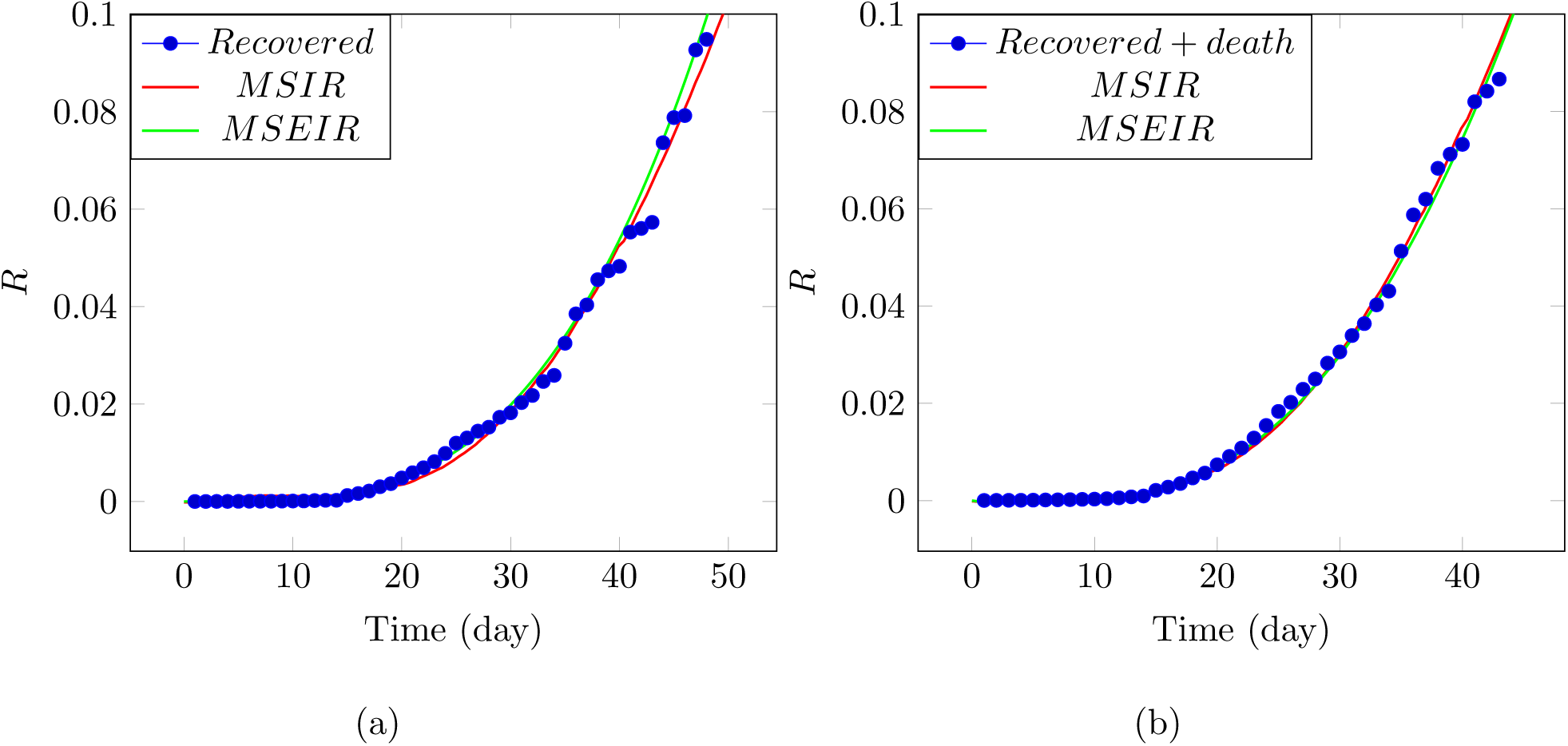
The fraction of the removed calculated from coronavirus disease (Covid-19) data https://en.wikipedia.org/wiki/2020_coronavirus_pandemic_in_the_United_States. These data are collected from the official reports from state health officials between March 12 to April 28, 2020, and fitted with MSIR and MSEIR models. The population size 1.5 million was used for calculating the fractions. (a) Only the recovered data are used for modeling; (b) Both the recovered and the death data are added together for modeling. *R*^2^ for both regressions are larger than 0.99.

**Figure 6:**
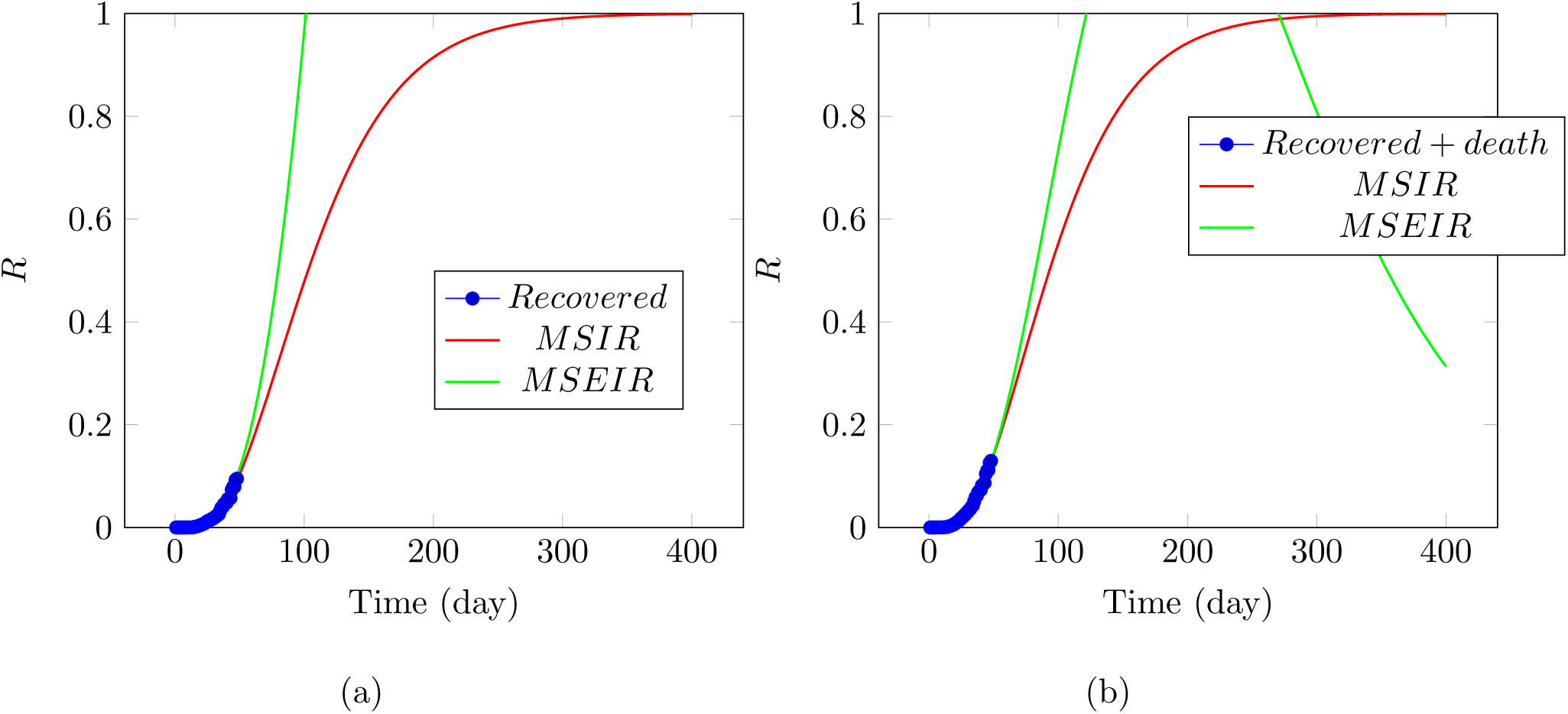
The fraction of the removed calculated from coronavirus disease (Covid-19) data https://en.wikipedia.org/wiki/2020_coronavirus_pandemic_in_the_United_States. These data are collected from the official reports from state health officials March 12 to April 28, 2020, and fitted with MSIR and MSEIR models for the recovered. The population size 1.5 million was used for calculating the fractions. (a) Only the recovered data are used for modeling; (b) Both the recovered and the death data are added together for modeling. *R*^2^ for both regressions are larger than 0.99.

**Figure 7:**
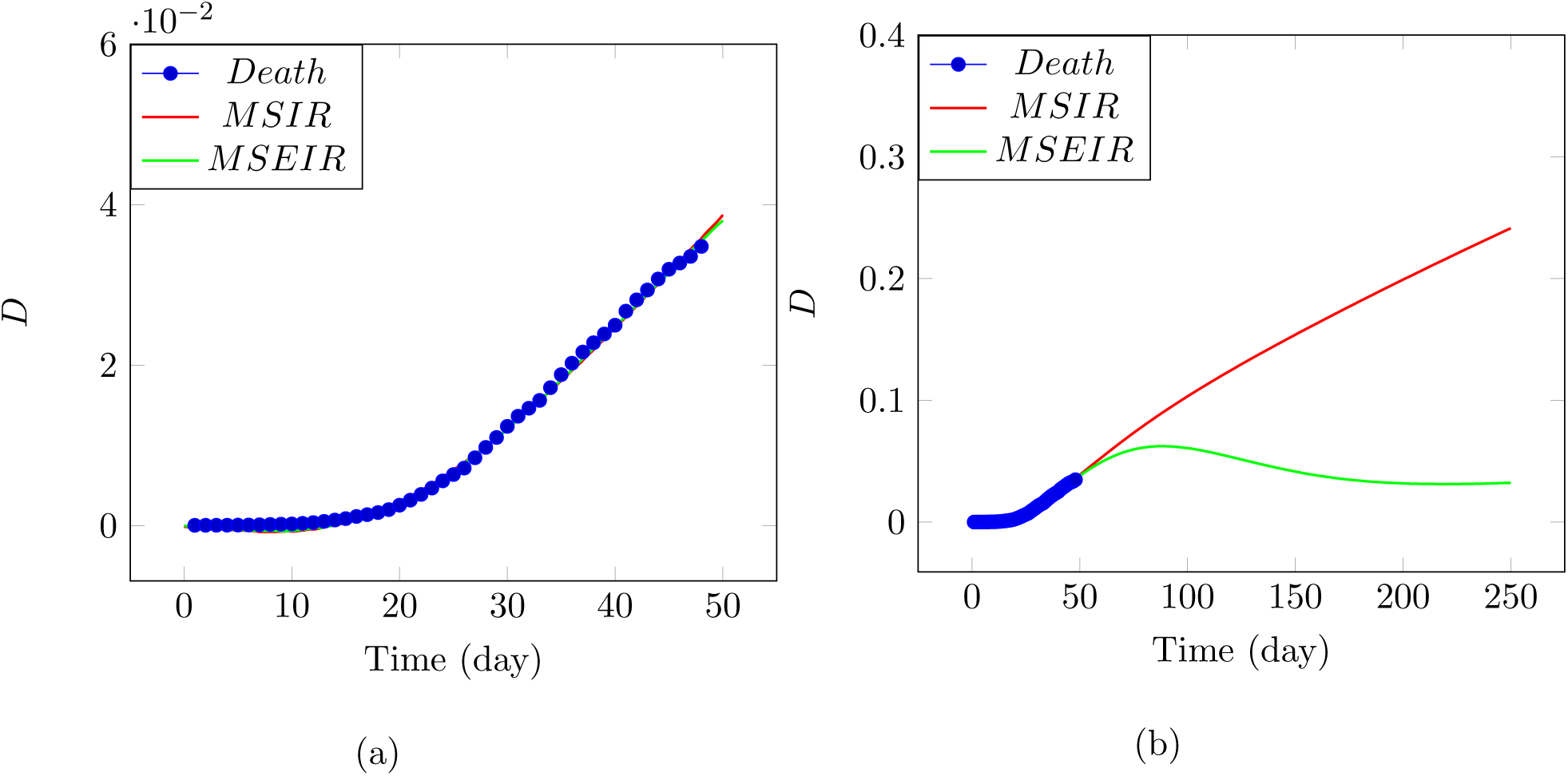
The fraction of the death calculated from coronavirus disease (Covid-19) data https://en.wikipedia.org/wiki/2020_coronavirus_pandemic_in_the_United_States. These data are collected from the official reports from state health officials between March 12 to April 28, 2020, and fitted with MSIR and MSEIR models for the removed. The population size 1.5 million was used for calculating the fractions. (a) small scale less than 50 days; (b) expanded to large scale. *R*^2^ for both regressions are larger than 0.99.

In summary, under the assumption that human movement energy may change with time during disease transmission period, we have modified the previous infection disease model formulated on the basis of the Eyring’s rate process theory and free volume concept^1^. Under such an approach, the model used in infectious disease transmission is consistent with the ones used in my previous publication for other systems. Treatment method is thus unified across different fields. Obtained equations fit the data very well with *R*^2^ large than 0.99 for all regressions. The predicted peak time, peak infected, death toll, and death rate is listed in Table I.

**Table I:** Peak time, peak infected, death toll estimated with MSIR and MSEIR models

| Model | Peak time from March 12, 2020(days) | Total infected expressed in fraction at peak time | Total infected expressed in population size at peak time (million) | Death toll at peak time (million) | Death rate % |
| --- | --- | --- | --- | --- | --- |
| MSIR | 115 | 0.66 | 1.98 | 0.18 | 9.06 |
| MSEIR | 64 | 0.36 | 1.08 | 0.08 | 7.37 |

## IV. DISCUSSION

The predictions made in this article are based on the data collected in USA and released from https://en.wikipedia.org/wiki/2020_coronavirus_pandemic_in_the_United_States. The main source of these data are from John Hopkins University https://www.arcgis.com/apps/opsdashboard/index.html#/bda7594740fd40299423467b48e9ecf6 and https://www.coronavirus.gov/. The differences among various sources are small, dependent on when the data are updated and released. The accuracy of predictions shouldn’t be affected by using different sources of data.

The integration of Eyring’s rate process theory and free volume concept has been demonstrated to work well for many multi-scale systems ranging from electrons to granular particles, even the universe^35^. This approach is therefore naturally applied to disease transmissions, as human movement and virus particle transmissions should follow same physical and chemical principles. Excellent fitting quality in term of *R*^2^ > 0.99 using the derived equations may indicate that the unified approach across multi-disciplinary areas does unveil fundamental operating mechanisms behind various phenomena.

The peak infected predicted with MSEIR model is 1.08 million, which will be reached earlier than May 16. Another parameter is death toll at peak time, 80,000 predicted with MSEIR model, which can be used as an indicator for peak time. We can see clearly from Figure 4 that the infected is more closely following the trend described with MSIR model rather than MSEIR model for last several days, implying that MSIR model may give a more plausible prediction.

An interesting and critical parameter introduced in this article is *β*, which defines the interaction level between human individuals during disease transmissions. *β* = 0.2 is found for the infected data regression and *β* = 0.01 is found for all other regressions, indicating that the interpersonal transmission is weak in USA at this moment and the isolation policy is working.

The theoretical framework proposed in this article can be applied to other countries and other transmission diseases, though the focus is put on covid-19 currently spreading in USA.

## V. CONCLUSION

With the argument that human movement energy is time dependent, we have modified our infectious disease transmission model proposed previously. The remaining formulation and structure of the model are unchanged: the infectious disease transmission process from the susceptible, to the exposed, the infected, and the removed in the end is continued to be considered as a sequential chemical reaction process, and the reaction rate at each step follows the Eyring’s rate process theory and free volume concept.

Obtained equations are employed to describe covid-19 outbreak currently ongoing in USA. Excellent fitting curves are obtained with *R*^2^ larger than 0.99 for all regressions including the infected, the removed (the recovered with and without the death), and the death toll alone. MSIR and MSEIR models give different predictions: MSIR model predicts that the infected will peak on July 7, 2020, with 1.98 million infected and 0.18 million death, while MSEIR model predicts that the infected will peak on May 16, 2020, with 1.08 million infected and 0.08 million death. The difference may be caused by the “exposed” category in MSEIR model, which may take a huge portion of the infected in MSIR model. The death rate predicted with MSIR model, is slightly higher than that predicted with MSEIR model, both are higher than the real number. The number of the infected are more closely following what is predicted with MSIR model for last several days.

The infection peak time is strongly dependent on the stretched exponential parameter *β*, which substantially amplifies peak heights. Large values of *β* mean that more people will be infected. For all regressions, the parameter *β* is less than 0.2, indicating that the interpersonal transmission is in a “weak” point and the “stay at home” isolation and travel restriction is working for preventing covid-19 from spreading.

The infection peak height is also dependent on the reaction rate constants such as *k*_2_, *k*_4_, *k*_5_. Small *k*_2_ means large peak heights, while large *k*_4_ and *k*_5_ lead to large peak heights, a substantially large number of infections.

## Data Availability

Available publicly

https://en.wikipedia.org/wiki/2020_coronavirus_pandemic_in_the_United_States

## Acknowledgments

The author sincerely appreciate Professor Yuanze Xu for his constructive feedback and comments for substantially improving the readability and rationality of this article.

